# The effect of care groups on infant and young child feeding, dietary diversity and WASH behaviours in rural Zimbabwe: A case-control study

**DOI:** 10.1101/2023.01.24.23284948

**Authors:** Tonderayi M. Matsungo, Faith Kamazizwa, Tafadzwa Mavhudzi, Starlet Makota, Blessing Kamunda, Calvin Matsinde, Dexter Chagwena, Kudzai Mukudoka, Prosper Chopera

## Abstract

**Background:** The care group model is a sustainable low cost community-based strategy to achieve nutrition related behaviour change and improve nutrition and diet quality indicators.

**Objective:** To evaluate the effect of the care groups on achieving positive infant and young child feeding, dietary diversity and WASH behaviours.

**Methods:** A case-control study utilised a mixed method approach targeting seven rural districts in Zimbabwe in June 2022. A structured questionnaire was used to collect data on IYCF, diet quality, WASH, and child morbidity. Binary logistic regression was used to evaluate the association between exposure (intervention) and outcome. Significance was at P<0.05.

**Results:** A total of 127 cases and 234 controls were enrolled. There was no significant difference between cases and controls on the prevalence of; diarrhoea (P=0.659), cough (P=0.191), and fever (P=0.916). No significant difference was observed in the proportion ever breastfed (P=0.609), diet quality-children with adequate dietary diversity score (CDDS) (P=0.606) across the two groups. However, the proportion of families with adequate Household Dietary Diversity Score (HDDS) (P=0.005) and minimum dietary diversity for women (MDD-W) (P=0.009) were significantly higher in cases than controls. Furthermore, the knowledge and practice of all promoted behaviours were significantly higher in the cases than in controls with exception of exclusive breastfeeding. Practice was significantly higher in cases compared to controls for: “Appropriate complementary feeding for children aged 6-24months” (P=0.001), “good nutrition for women of childbearing age” (P=0.001), “production and consumption of diverse nutritious food” (P=0.001) and “production and consumption of biofortified crops” (P=0.001).

**Conclusions:** The current results adds to the body of evidence showing that care groups are effective for achieving sustainable nutrition and WASH related behaviour change in a low-income setting. Overall, care groups should be implemented to scale ‘coverage” and integrated into existing community nutrition programs for maximum impact.

**Key Messages:**

- The current results add onto the limited evidence showing the potential of care group approach towards improved IYCF, dietary diversity, WASH behaviours and practices in low-income settings. However, this approach should be implemented to scale and not in isolated pockets of the community to ensure positive impact on nutrition indicators.
- The care group SBCC strategy led to improved knowledge and selected practices. However, there was no significant impact on child morbidity indicators and child diet quality indicators.
- The care group approach should be integrated into community-based nutrition programming guided by the key lessons learned from pilot districts.
- Emphasis should be on how improved knowledge and practices can translate to improved child IYCF indicators.

## Introduction

The burden of malnutrition has remained a public health concern for Zimbabwe. Prevalence of stunting is still high at 29.4% ^1^. Major malnutrition drivers in Zimbabwe have been intensified by economic, social, environmental and political shocks and stresses, many of which have devastating effects on vulnerable populations ^2^. When faced with challenges, communities often cope by striving towards food quantity as opposed to food quality, thereby compromising their nutrition well-being ^3^. The majority poor have resultantly been exposed to high levels of food insecurity, malnutrition, micronutrient deficiencies, poor service delivery, as well as compromised access and utilisation of available foods – all these possibly contributing towards high stunting levels. Stunting is the leading form of malnutrition among children younger than 5 years worldwide ^4^. In Zimbabwe, 1 in 3 children under five years is affected by stunting, 29.4% ^1,5^. Although the causes are multifactorial and diverse, recuring infections and poor infant and young child feeding (IYCF) practices are the preeminent factors associated with stunting. In Zimbabwe, improving diet quality and reducing the prevalence of stunting has been a challenge^6^. Therefore, there is need to focus interventions on a multi-sectorial approach to effectively address stunting and the underlying determinants.

The care group approach is an innovative community-based strategy, used to improve nutrition behaviour change in a large population while maintaining low cost and sustainability ^7^. This approach has strengthened communities’ resilience to repeated shocks though in some implementation sites progress has lagged behind despite large scale implementation ^8^. The care group approach is a promising model to promote behaviour change towards improved IYCF practices in low income settings, where health workers have heavy workloads ^9^. Few studies done in Zimbabwe evaluating overall effectiveness of the care group approach have found that this approach has improved IYCF, WASH behaviours and practices and has opportunity to include various age groups such as adolescents ^9–12^. No study has been done to evaluate the effectiveness of the care group SBCC strategy on selected behaviours, nutrition and health outcomes. The objective of this study were therefore to investigate the effectiveness of the care group model SBCC strategy in influencing behaviour change of promoted nutrition and health practices.

## Methods

### Study setting

This study was conducted in seven randomly selected districts participating in the Zimbabwe Resilience Building Fund (ZRBF) programme in Chiredzi (Masvingo), Beitbridge (Matabeleland North), Mutoko (Mashonaland East), Kariba (Mashonaland West), Mberengwa (Midlands), Bubi (Matabeleland North), and Insiza (Matabeleland South) (**Figure 1**). These districts -are in agro-ecological regions 4 and 5 and were participating in the nutrition sensitive agriculture and resilience building programme (ZRBF) and in addition the beneficiaries also participated in the nutrition focused SBCC care groups.

**Figure 1:**
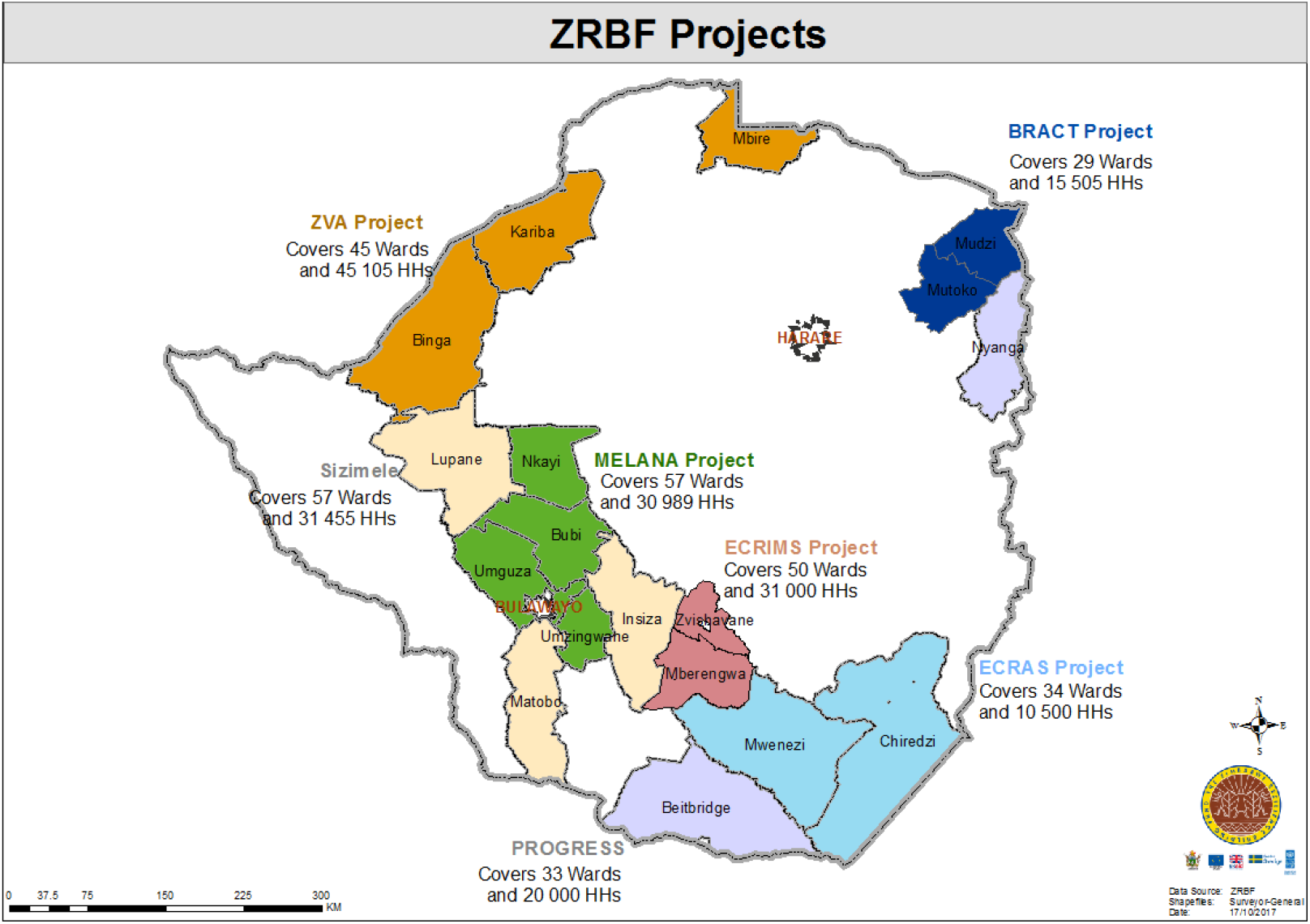
Map showing the ZRBF project implementation districts in Zimbabwe

### The care group approach (CGA)

The target respondents for the quantitative questionnaire were ZRBF beneficiaries particularly women participating in care groups. CGA is a nutritional behavioural approach adopted by the Government of Zimbabwe, implemented by Nutrition Action Zimbabwe (NAZ) a local NGO as an effort to mainstream nutrition into ZRBF which was primarily an agriculture and livelihoods improvement multisectoral intervention. The CGA is an internationally recognized strategy and a series of social and behavioural change interventions aimed at reducing malnutrition and child mortality. CGA was first developed by World Relief in 1995 during a child survival project in the Gija and Mabarane districts of Gaza Province, Mozambique ^7^. It has proven to be a convergent platform to discuss and practice both nutrition specific and nutrition sensitive interventions ^9^.

In ZRBF, the CGA was constructed based on global guidance, where volunteers (lead mothers) motivate mothers (neighbour women/ beneficiaries) to adopt key nutrition and health behaviours ^7^. Lead mothers meet in groups with promoter once a month to learn new health and nutrition promotion messages. The main component consists of mothers teaching each other about these promoted behaviours in a network of households. The lead mothers have a manageable workload of up to 15 households or neighbour women constituting a care group, where they meet at least once a month for support and supervision. The detailed description of CGA model in the Zimbabwean context has been described previously by Macheka et al., (2021). The ZRBF care groups promoted seven behaviours; (1) exclusive breastfeeding for children from birth to 6 months (2) Offer children aged 6-23 months, timely, adequate and diverse complementary feeding with continued breastfeeding up to 2yrs and beyond (At least 4 of the 8 food groups). (3) Consumption of diverse foods by women of childbearing age (15-49 years) (4) Mothers of children 0-59 months wash their hands at the five critical times. (5) Food preservation of at least four types of foods from the recent farming season for household consumption later in the year. (6) Household production and consumption of diverse nutritious foods including Neglected Underutilized Foods (e.g., small grains and wildly harvested vegetables) at least 3 times per week). (7) Purchase of nutritious, diverse foods including animal source foods and legumes utilizing agricultural income.

### Study design and sampling

The study utilized an unmatched case control design. Seven districts were randomly selected out of the possible 18 implementing districts (cases). The controls were also recruited from the same districts but in wards that were not implementing ZRBF activities with bias on the same province thus reducing confounding errors. Individuals/households who participated in the care groups were enrolled as cases. While individuals/households who did not get the opportunity to be in care groups were enrolled as controls. We used a ratio of cases to controls of 1:2 respectively according to methods described by Sullivan & Soe (2022)^13^. We used 80% power to obtain a sample size of 411 for households (137 cases & 274 controls) distributed across the seven study districts.

### Data Collection and tools

The study utilised a mixed methods approach were both quantitative (structured questionnaire) and qualitative data (FGD and KIIs) was collected from participants for the purposes of triangulation.

#### Structured questionnaire (Android based)

A validated questionnaire adapted from the ZIMVAC survey tool was used^2^. The questionnaire had the following sections: demographics, socioeconomic information, participation in care groups, agricultural, water sanitation and hygiene (WASH), infant and young child feeding (IYCF) practices. The questionnaire assessed the seven ZRBF promoted behaviours in a series of yes or no questions. The questionnaire was converted into an electronic version and uploaded on to android tablets using the Kobo Collect Toolbox application (Kobo Inc, Cambridge, UK). All enumerators were given tablets with the electronic questionnaire. The questionnaire was administered by trained enumerators who used the local language. The following diet quality indicators, household, women and child dietary diversity were assessed;

#### Household dietary diversity score (HDDS)

Home diet diversity is used to measure the quality of home diets, especially macro- and micronutrients. Food intake data were collected using the 24-hour recall method. This consisted of accurately recalling, describing and/or computing food and drink consumed in her 24 hours or the day before the interview ^14^… The latter mentioned twelve food groups were used to calculate household dietary diversity scores; (1) Cereals, (2) Roots and tubers, (3) Vegetables, (4) Fruits, (5) Meat, poultry, and offal, (6) Eggs, (7) Fish and seafood, (8) Pulse, legumes, and nuts, (9) Milk and milk products, (10) Oil/ fats, (11) Sugar/ honey and (12) Miscellaneous. Households that have consumed one of the above mentioned food groups within the last 24 hours receive a score of 1. The HDDS variable is calculated per household. The range of values for this variable is zero (0) to twelve (12). ^15^.

#### Minimum Dietary Diversity women (MDD-W)

Dietary diversity thresholds for women were measured according to the FAO guidelines for measuring dietary diversity thresholds for women ^16^. This indicator measures the adequacy of micronutrients in women’s diets at the population level. Women of childbearing age (15–49 years) recorded all food consumed during the previous day or night (last 24 hours) at home or on the go. Foods were assigned to 10 recommended food groups: (1) Grains, roots, and tubers, (2) Pulses, (3) Nuts and seeds, (4) Dairy, (5) Meat, poultry, and fish (6) Eggs (7) Dark leafy greens and vegetables (8) Other Vitamin A-rich fruits and Vegetables (9) Other vegetables (10) Other fruits. The threshold for adequacy was defined as 5 or more food groups.

#### Child dietary diversity score (CDDS)

Minimum dietary diversity child was determined according to the new IYCF indicator guidelines ^17^. Breastmilk was included as the 8th food group and the cut off for adequacy was at least 5 out of 8 food groups, including breastmilk. The food groups included therefore were; breastmilk, grains roots and tubers, legumes and nuts, dairy products, flesh foods, eggs, vitamin A rich fruits and vegetables, and other fruits and vegetables.

#### Child morbidity

Prevalence of the 3 most common childhood ailments, diarrhoea, cough and fever in the study children was assessed by research assistants by interviewing the caregivers. The recall period for all symptoms was 2 weeks. Prevalence was determined as number of reported events over total number of recalls.

### Qualitative data collection

#### Focus Group Discussions (FGDs)

The FGDs consisted of 6-12 people, 1 per district (7 FGDs). Women of reproductive age (15-49 years) and beneficiaries of the care groups were randomly selected to participate in FGDs by the Enumerators. FGD sites were conveniently selected by the enumerators. A focus group guide was developed specifically for the study. The FGD were held up to the point where consecutive discussions revealed no additional data (saturation).

#### Key Informant Interviews (KIIs)

A total of 37 key informants were purposively selected and they included government officials (Agritex officers, district nutritionists, nutrition assistants, sisters in charge and village health workers), program officers (ZRBF, NAZ) and community leaders (village heads), lead farmers and lead mothers.

### Data analysis

All quantitative data was exported to SPSS version 24 (IBM Inc, NY, USA) from Kobo Collect data base. The frequency of each measured continuous outcome in each of the two groups (cases and controls) was calculated. Case control studies typically produce the odds ratio to measure strength of association between the intervention (exposure) and outcome. Variables with odds ratio greater than 3 were included in a multivariate logistic regression model (by enter method) to find adjusted odds ratio. Statistical significance was set at P<0.05. Qualitative data was analysed in Excel using content analysis to establish emerging themes. The most widely adopted six-step framework for conducting thematic analysis was employed. This consists of i) researcher familiarisation with the data ii) generating initial codes, iii) searching for themes iv) reviewing themes v) defining and naming themes vi) producing the report^18^.

## Results

### Socio-demographics

The study successfully enrolled 127 cases and 234 controls from seven rural districts in Zimbabwe. Majority of the participants were in the age group 25-64 years, cases (75%) and control (64%) respectively (**Table 2**). The household heads were mostly males, 79.3% in cases and 72% in controls and most of these were married (cases, 84.8% and 72.3% controls). Farming was the major economic activity in the study districts (cases, 79.3% and 64.6% controls). While the most frequent level of education was primary level in cases (45.7%) and secondary education in controls (45.7%).

**Table 1:**
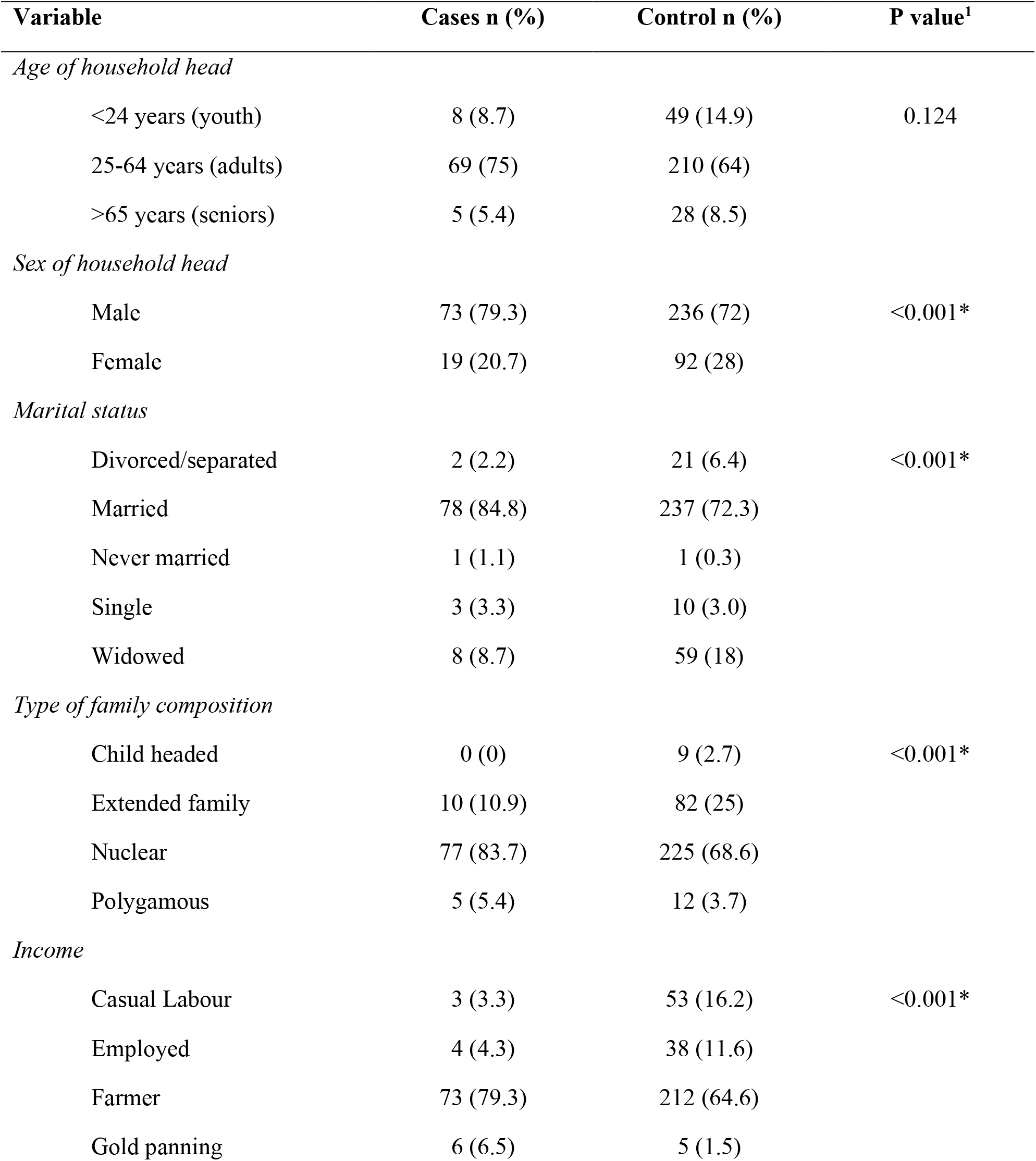
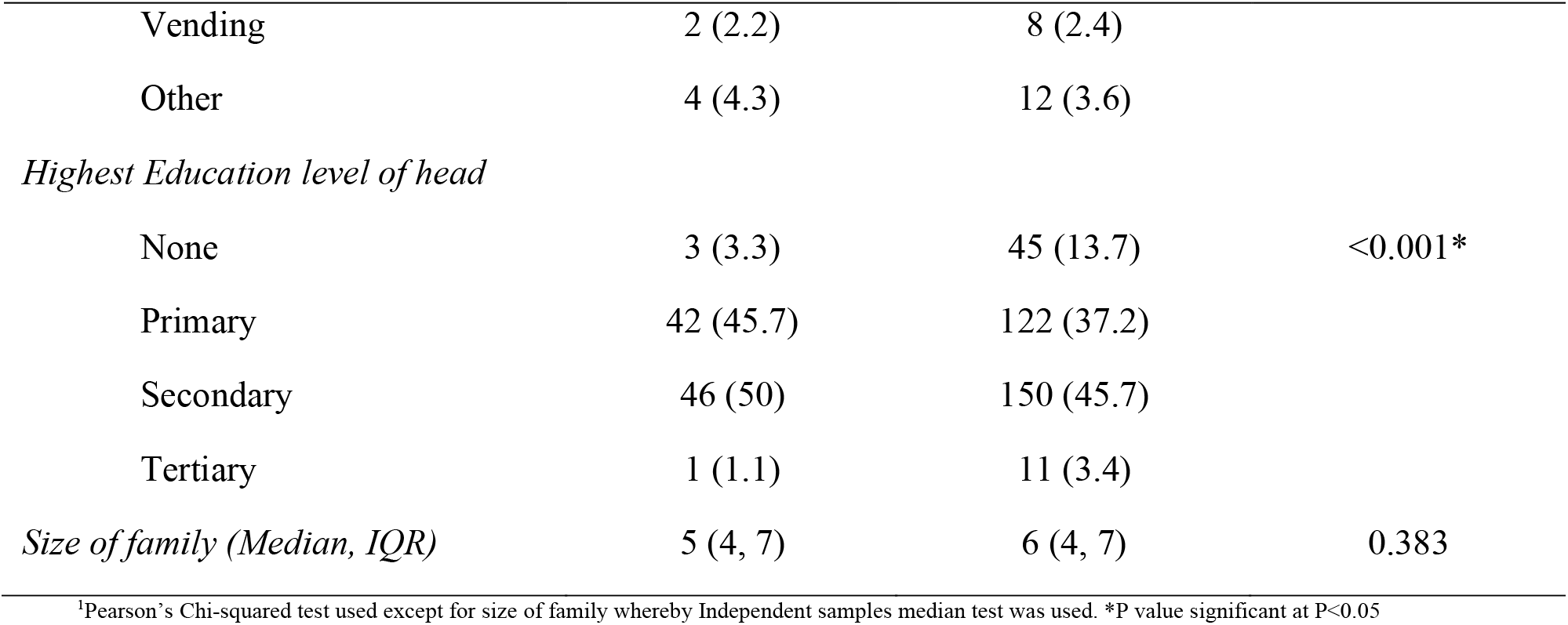
Sociodemographic characteristics of study households

| Variable | Cases n (%) | Control n (%) | P value<sup>1</sup> |
| --- | --- | --- | --- |
| *Age of household head* |  |  |  |
| <24 years (youth) | 8 (8.7) | 49 (14.9) | 0.124 |
| 25-64 years (adults) | 69 (75) | 210 (64) |  |
| >65 years (seniors) | 5 (5.4) | 28 (8.5) |  |
| *Sex of household head* |  |  |  |
| Male | 73 (79.3) | 236 (72) | <0.001* |
| Female | 19 (20.7) | 92 (28) |  |
| *Marital status* |  |  |  |
| Divorced/separated | 2 (2.2) | 21 (6.4) | <0.001* |
| Married | 78 (84.8) | 237 (72.3) |  |
| Never married | 1 (1.1) | 1 (0.3) |  |
| Single | 3 (3.3) | 10 (3.0) |  |
| Widowed | 8 (8.7) | 59 (18) |  |
| *Type of family composition* |  |  |  |
| Child headed | 0 (0) | 9 (2.7) | <0.001* |
| Extended family | 10 (10.9) | 82 (25) |  |
| Nuclear | 77 (83.7) | 225 (68.6) |  |
| Polygamous | 5 (5.4) | 12 (3.7) |  |
| *Income* |  |  |  |
| Casual Labour | 3 (3.3) | 53 (16.2) | <0.001* |
| Employed | 4 (4.3) | 38 (11.6) |  |
| Farmer | 73 (79.3) | 212 (64.6) |  |
| Gold panning | 6 (6.5) | 5 (1.5) |  |

| **Variable** | **Cases n (%)** | **Control n (%)** | **P value<sup>1</sup>** |
| --- | --- | --- | --- |
| Vending | 2 (2.2) | 8 (2.4) |  |
| Other | 4 (4.3) | 12 (3.6) |  |
| *Highest Education level of head* |  |  |  |
| None | 3 (3.3) | 45 (13.7) | <0.001* |
| Primary | 42 (45.7) | 122 (37.2) |  |
| Secondary | 46 (50) | 150 (45.7) |  |
| Tertiary | 1 (1.1) | 11 (3.4) |  |
| *Size of family (Median, IQR)* | 5 (4, 7) | 6 (4, 7) | 0.383 |

<sup>1</sup>Pearson's Chi-squared test used except for size of family whereby Independent samples median test was used. \*P value significant at P<0.05

**Table 2:** Child Indicators of diet quality and disease prevalence

| Variable | Cases |  | Controls |  | Odds Ratio<sup>1</sup> | 95%CI | P value<sup>1</sup> |
| --- | --- | --- | --- | --- | --- | --- | --- |
|  | n | % | n | % |  |  |  |
| *Select IYCF indicators* |  |  |  |  |  |  |  |
| Ever breastfed (Yes) | 35 | 97.2 | 58 | 95.1 | 1.81 | 0.18-18.09 | 0.609 |
| (No) | 1 | 2.8 | 3 | 4.9 |  |  |  |
| *Child Dietary Diversity Score (CDDS)* |  |  |  |  |  |  |  |
| <5 Food groups<sup>2</sup> | 26 | 72.3 | 41 | 66.2 | 0.79 | 0.32-1.95 | 0.606 |
| > 5 Food groups | 10 | 27.7 | 21 | 33.8 |  |  |  |
| *Disease prevalence* |  |  |  |  |  |  |  |
| Diarrhoea (Yes) | 8 | 22.2 | 16 | 26.2 | 0.80 | 0.30-2.12 | 0.659 |
| (No) | 28 | 77.8 | 45 | 73.8 |  |  |  |
| Cough (Yes) | 26 | 72 | 36 | 59.1 | 1.81 | 0.74-4.40 | 0.191 |
| (No) | 10 | 28 | 25 | 40.9 |  |  |  |
| Fever (Yes) | 8 | 22.3 | 13 | 21.4 | 1.06 | 0.39-2.86 | 0.916 |
| (No) | 28 | 77.7 | 48 | 78.6 |  |  |  |

<sup>1</sup>Binary Logistic regression Odds ratio (OR), <sup>2</sup>New 8 food groups for children calculation was used (Breastmilk is the new standalone group)

### Child indicators of diet quality and morbidity

The results (**Table 2**) showed both positive and negative trends. Although not statistically significant we observed lower prevalence of diarrhoea (P=0.659), cough (P=0.191), and fever (P=0.916), in the cases group. While the negative trends included, lower proportion of ever breastfed in the cases group (P=0.609), and lower proportion of children with adequate dietary diversity score (CDDS) in the cases group (P=0.606), an indication of poorer diet quality in cases than controls. However, these trends were all not significant.

### Perceptions of the promoted behaviours

Based on the qualitative data (FGDs and KIIs), participants think that it is necessary to adopt the seven promoted behaviours so as to have well-nourished children and families and to maintain cleanliness at all times.

> *“If we implement these behaviours then our communities will be better with less children getting sick“*
>
> *>>>FGD #1*
>
> *“VSL activities are helping improve our livelihoods. Improved household sanitation, rubbish t pits, tippy taps, pot racks and upgradeable pit latrines). “*
>
> *>>> FGD #2*
>
> *“There has been increased hand washing, we are now eating balanced diets and our children are growing well.“*
>
> *>>> FGD #3*
>
> *“Reduced diarrhoea cases, no more open defecation“*
>
> *>>> FGD #4*

Diversification of diets was appreciated by the cases and they were able to incorporate different available foods into their children’s diets as shown below:

> *“I can now make a four-star diet for my 18months old baby without buying anything“*
>
> *>>> FGD #5*
>
> *“There were improvements in infant feeding practice. For example, before the programme children aged 6 to 23 months were given only sour porridge*“
>
> Lead Mother, District #1
>
> “*Direct statistics are difficulty to find but our observation now are that child nutrition has changed for the better especially foods given for complementary feeding*”
>
> Ministry of Health official, District #2

### Household and adult indicators WASH, diet quality, promoted behaviours

The results in **Table 3** revealed that knowledge and practice of all promoted behaviours except exclusive breastfeeding were significantly higher in the cases. Practice was significantly higher in cases compared to controls for: “Appropriate complementary feeding for children aged 6-24months” (P=0.001), “good nutrition for women of childbearing age” (P=0.001), “production and consumption of diverse nutritious food” (P=0.001) and “production and consumption of biofortified crops” (P=0.001). These are positive findings are implying the possible impact of the care group approach in promoting nutrition and health related behaviours. However, the specific results of these indicators are presented in next section.

**Table 3:**
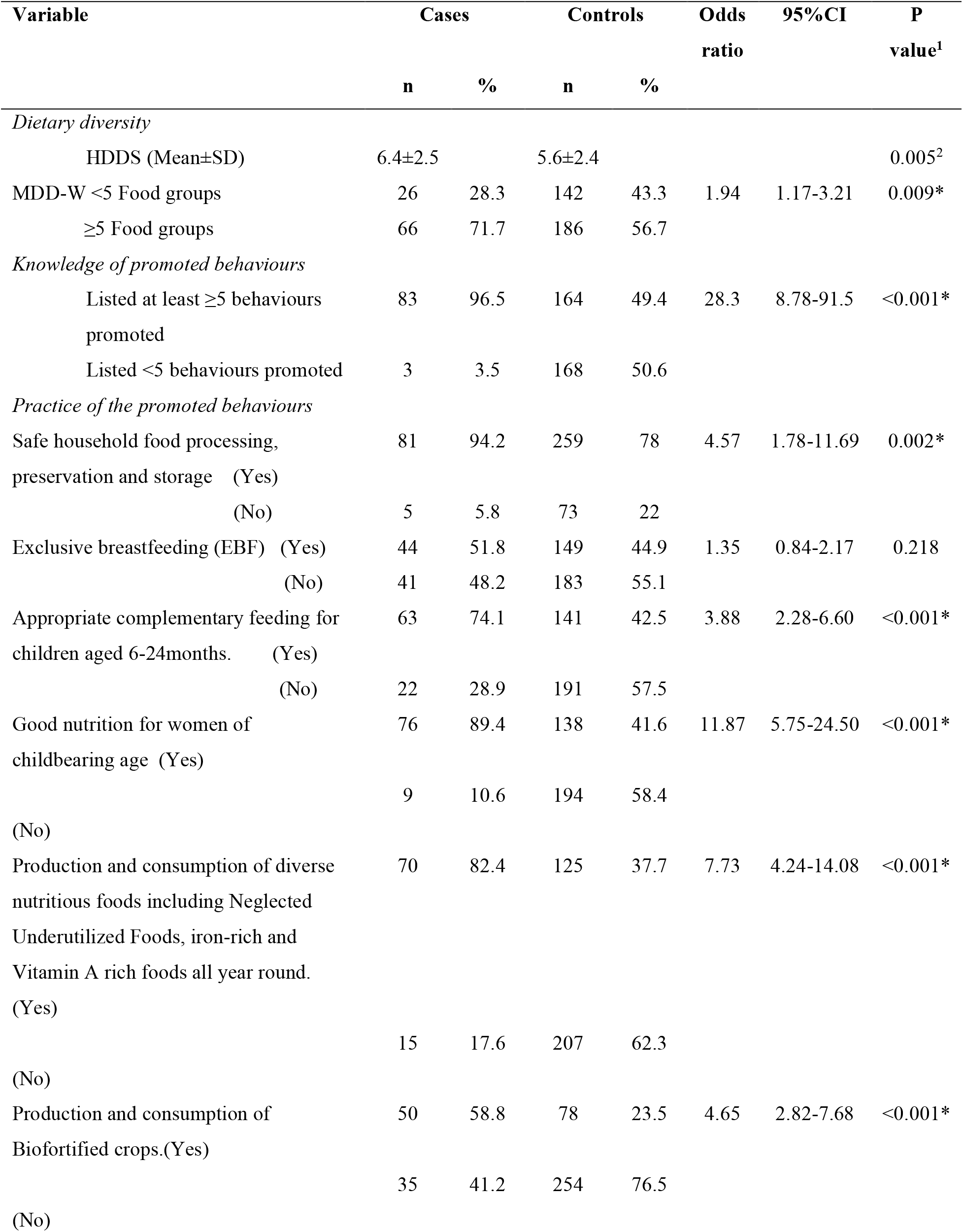
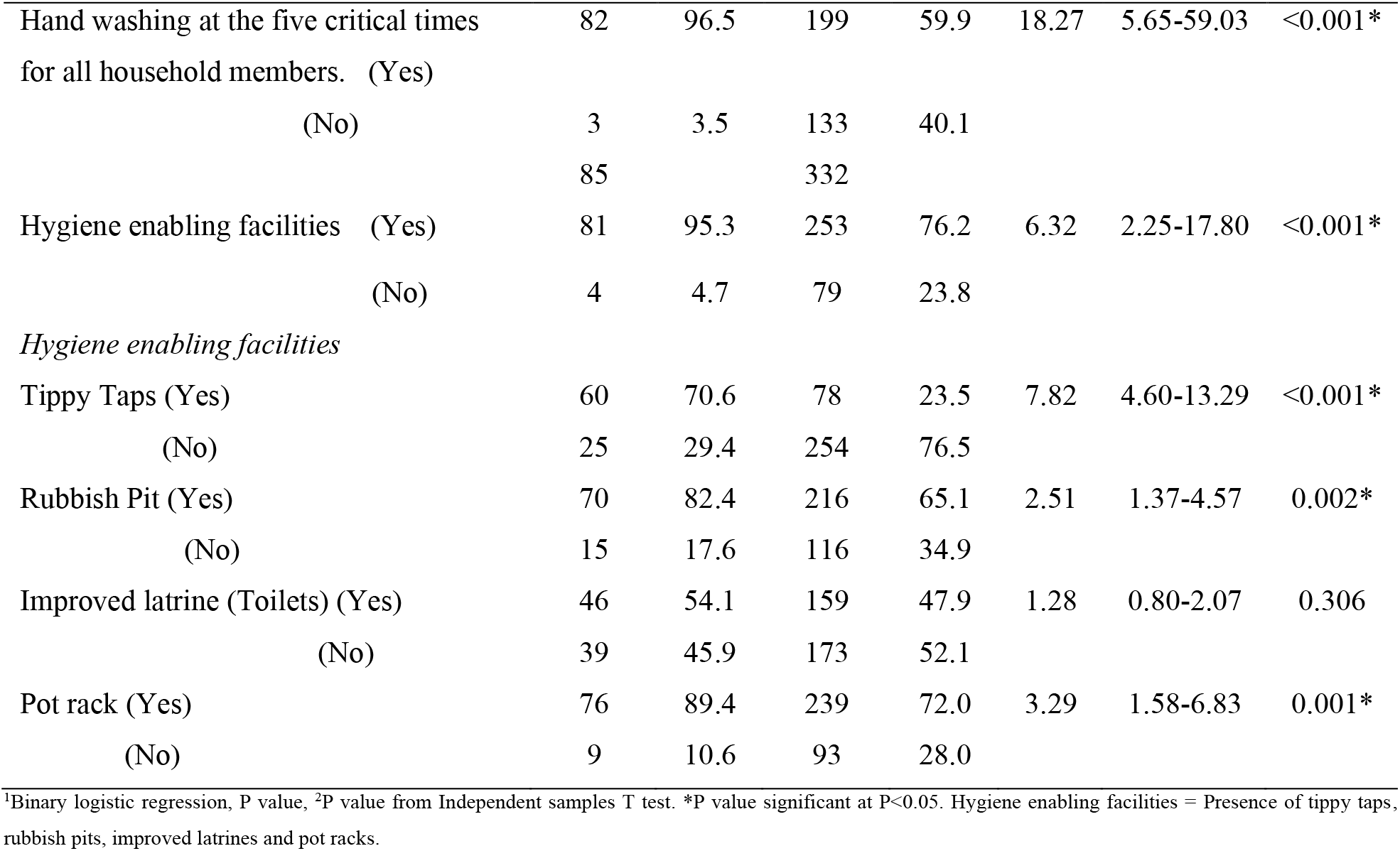
Household and adult indicators WASH, diet quality, promoted behaviours

#### Food security and dietary diversity

Household dietary diversity scores (HDDS) and minimum dietary diversity for women (MDD-W) were significantly higher in cases (96.5%) compared to controls (49.4%) (P=0.005). This finding corresponded with qualitative data recorded from the KIIs and FGDs. For instance, majority of the KII mentioned the increased production of diversified crops (including small grains) as well as small livestock (chicken, fisheries) production to ensure availability of four-star diets. Adoption of production and consumption of biofortified crops especially NUA 45 beans and orange sweet potatoes was also highlighted. The care groups have resulted in knowledge exchange especially on types of crops that can be grown all year round with high nutritive values

> *“The changes brought about by care groups are quite positive”*
>
> Lead Mother, District #2
>
> “*Behaviours encourages production and consumption of diversified diets thus resulting in good nutrition for very community member, and community becomes self-sufficient and resilient“*
>
> *>>>FGD #6*
>
> *“Using locally available foods saves money and we can use it for other basic commodities“*
>
> *>>>FGD #7*

#### Knowledge of promoted behaviours

Our findings showed that knowledge of the seven promoted behaviours was also higher in the cases compared to controls. The proportion of participants who remembered at least five behaviours was higher in cases (96%) compared to controls (49.4%) (P<0.001). Key informants strongly recommended that the consortia should promote all the 7 behaviours in all participating districts.

> “*Some of the behaviours were not promoted in our district, only IYCF indicators were promoted*“’
>
> *>>>FGD #5*

#### Practice of the promoted behaviours

One of the FGD participants highlighted that

> *“Exclusive breastfeeding is difficult to practice in our churches we get holy water to protect the child from bad spirits”*
>
> *>>>FGD #1*
>
> *“Some religious groups shy away from taking part in health-related activities, hindering program impact. In addition, we have some bad cultural beliefs that hinder adoption of IYCF practices in our communities”*
>
> Ministry of Health official, District #3
>
> *“The community is accepting the program, though some cultural beliefs affect adoption of certain behaviours especially on breastfeeding”*
>
> Lead Mother, District #3

#### Hygiene enabling facilities

Hygiene enabling facilities were significantly higher in ZRBF wards with the exception of improved latrines (P=0.306). Similar to the quantitative data there were field observations of pot racks, rubbish pits, tippy taps and advanced latrines in some households.

> *“I taught them about the tippy taps, rubbish pits, pot rack, use of toilets, even temporary toilets’*
>
> >>> Lead Mother, District #4

The proportion of cases *“practicing hand washing at the five critical times for all household members”* was significantly higher compared to controls (P<0.001).

## Discussion

The study sought to evaluate the contribution of the care group approach as an SBCC strategy to improved nutrition and health related behaviours. The results revealed that care groups are a good entry point for mainstreaming of nutrition within resilience and agriculture-based interventions. Integrating nutrition into new and existing and programs is critical to addressing the urgent and pervasive problem of malnutrition in low-income communities We observed that through the care groups lead mothers motivated their neighbours (pregnant and lactating) to adopt positive nutrition and health behaviours with support from lower level nutrition governance structures ^7^. Thus the results demonstrate the value of promoting nutrition education and behaviour change utilising the care group or mother led groups and support the call to integrate this approach into community nutrition programmes.

### Child morbidities

This study showed both positive and negative trends. On the positive side, there was relatively lower prevalence of diarrhoea, cough and fever in the participating wards (cases). A cross-sectional baseline survey carried out by Hillow in Ethiopia also showed a significant decrease in reported cases of child diarrhoea ^19^. Similar success with the care group model has been observed in other countries such as Cambodia, Mozambique, Malawi, Kenya and Rwanda with decline in child mortality, and improved IYCF indicators^7^. In addition, a study in Uganda that assessed the contribution of volunteer community health workers on child morbidity reported a decreased child morbidity^20^. Therefore, the prevention and management of child morbidities depends on several individual, household and community level factors. The care groups offer a comprehensive approach to tackle these complex factors.

### Indicators of diet quality for children

The proportion of children with adequate dietary diversity score (CDDS) was lower in participating wards an indication of poorer child diet quality in cases than controls. However, these trends were all not significant an indication that the relationship between exposure and outcome was by chance. According to the qualitative data diversification of diets was appreciated by incorporating different foods. These are reported behaviour rates and may not reflect actual changes. Overall, the NAZ-ZRBF intervention appears to be effective on WASH variables and overall disease prevalence but does not appear to be effective on children’s diet quality.

### Food security and dietary diversity

Household dietary diversity scores (HDDS) and minimum dietary diversity for women (MDD-W) were significantly higher in cases compared to controls .Findings from FGDs with the neighbour women revealed that mothers felt that the ZRBF nutrition intervention brought significant improvements in terms of feeding habits of the communities as well as improvements in consumption of diversified diets as they now had nutrition home gardens. Household production of small grains like sesame, sorghum, finger millet and seeds were observed. Our findings reflect a positive impact of the care group approach on increased household food supply and agree with earlier findings from Cambodia, Mozambique, Malawi, Kenya and Rwanda ^7^. It is however concerning that the improved diet quality at the household level is not translating to improved child diet quality. This could be as a result of poor intra household food distribution that disadvantages children within the African context possibly stemming from traditional and cultural beliefs. Therefore, future studies that investigate factors affecting intra household food distribution, and at large social determinants of children’s dietary intake are required.

### Knowledge of promoted behaviours

Our findings showed that knowledge of the seven promoted behaviours was higher in the cases compared to controls. The proportion of participants who remembered at least five behaviours was higher in the participating wards compared to the non-participating wards. This is a positive finding showing positive impact of the care group approach on nutrition knowledge among pregnant and lactating mothers. Our findings are similar to findings by Macheka et al., (2022) who reported a relatively high knowledge of the nutritional behaviours by participating households^21^. In the use of the case control design in the current study could explain the observed differences with Macheka et al., (2022) results. However, in the latter studies knowledge was also high in non-participating households and attributed this to spill over effects of the care group trainings. In overall both studies reveal that participation in care groups has a positive effect on nutrition knowledge.

### Practice of the promoted behaviours

Our findings revealed that practice of all promoted behaviours were significantly higher in the participating wards except exclusive breastfeeding. Exclusive breastfeeding is a behaviour promoted by all in the health sector in Zimbabwe as the country is a signatory to the International Code of Marketing of Breastmilk Substitutes. This behaviour is expected to be high and similar across cases and controls. However in contrary to our findings a mid-term review of the Care Group Model in Zimbabwe revealed that there was a shift in practice towards the adoption of all the new behaviours promoted through the care group model including exclusive breastfeeding ^9^. It could be that different methods have been used to assess breastfeeding practices hence the divergent results. It could also be that behaviour change takes time as such comparison of mid-term and end term review may not be appropriate. Interestingly, our results agree with Macheka et al., (2022) and Gomora et al., (2019) who reported that participation in care groups resulted adoption of positive nutrition behaviours.

### Hygiene enabling facilities

In this study hygiene enabling facilities were significantly higher in the participating wards. There were field observations of pot racks, rubbish pits, tippy taps and pit latrines in some households. This is similar to findings by Gomora et al., (2019) where many care group clients had adopted the new behaviours being promoted and this was evidenced by construction of sanitation and hygiene enabling facilities, redesigning of a new and more durable tippy tap, rubbish pits and pot racks at their households ^9^. Macheka et al., (2022) also observed positive impact of care groups on the adoption of WASH behaviours in rural communities in Zimbabwe ^21^..

### Strengths and limitations of the study

Our study had several strengths. We used a case control design. Though this design requires a smaller sample than does for example a cohort study it can be used to detect the same level of increased risk. We also combined quantitative and qualitative methods. This makes the study design and findings more robust. Whilst in a quantitative design the relationships between variables are measurable, qualitative design helps elucidate drivers of human behaviours in much more depth and detail. Further parameters such as diet quality were assessed using validated methodology^15–17^. One weakness of our study is that some districts like Chiredzi had been on the programme for longer time than the other districts thus possibly introducing selection bias. Although, case-control studies may prove an association they do not demonstrate causation.

## Conclusions

The care group SBCC strategy led to improved knowledge, breastfeeding, complementary feeding and WASH behaviours. However, there was no significant impact on child morbidity indicators and diet quality. Overall, our results add on to the body of knowledge demonstrating the effectiveness of the care group approach as a tool for achieving behaviour change in low-income settings. The, Government of Zimbabwe should consider nationwide integration of the care group approach into community nutrition and/or resilience building programs guided by the lessons learned. Most importantly, these results should inform the review of the national care group guidelines for Zimbabwe.

## Data Availability

The anonymised datasets used and/or analysed during the current study are available from the corresponding author on reasonable request.

## Acknowledgements

We are very grateful to the participants for agreeing to participate in this study. Great thank you also to the key informants and mothers who participated in the group discussions for sharing their valuable perspectives. We are grateful to ZRBF consortia staff for feedback on the protocol and supporting the data collection phase.

